# Inhibition of HIF-2α Pathway as a Potential Therapeutic Strategy for Endothelial Dysfunction in Post-COVID Syndrome

**DOI:** 10.1101/2024.09.10.24313403

**Authors:** Andrea Ribeiro, Timon Kuchler, Maciej Lech, Javier Carbajo-Lozoya, Kristina Adorjan, Hans Christian Stubbe, Martina Seifert, Anna Wöhnle, Veronika Kesseler, Johanna Negele, Uwe Heemann, Christoph Schmaderer

## Abstract

**Background:** SARS-CoV-2 infection may lead to Post-COVID Syndrome (PCS), characterized by debilitating symptoms like persistent fatigue, cardiovascular symptoms, and cognitive dysfunction. Persistent endothelial dysfunction (ED) is a potential driver of ongoing symptoms. Yet, the underlying biological mechanisms remain unclear.

**Methods:** In this prospective observational study, we characterized 41 PCS patients and 24 healthy controls (HC, matched out of n = 204, recruited before the pandemic) and investigated the effect of SARS-CoV-2 Spike protein 1 (S1) and plasma from PCS patients on human retinal endothelial cells (HREC).

**Results:** Plasma samples from PCS patients exhibited significantly elevated erythropoietin, VEGF and MCP-1 alongside decreased IL-6 levels compared to HC. Low Haemoglobin and Haematocrit were negatively associated with PCS severity. VEGF levels were positively correlated with Anti-S1 IgG levels in patients and upregulated on mRNA level in HREC exposed to S1. Additionally, S1 exposure promoted ROS production and transiently activated HIF-1α in HREC. Persistent activation of HIF-2α by S1 led to disrupted endothelial integrity. HREC exposed to plasma from severely affected PCS patients showed increased ROS and compromised barrier function. Treatment with Belzutifan, a HIF-2α inhibitor, restored barrier integrity in HREC exposed to S1 or PCS-plasma.

**Conclusion:** These findings suggest that HIF-2α-mediated ED in PCS might be a potential therapeutical target for Belzutifan.

**Trial registration:** URL: https://www.clinicaltrials.gov; Unique identifier: NCT05635552

**Novelty and significance:** *What Is Known?:*

- Endothelial dysfunction (ED) is a consequence of acute SARS-CoV-2 infection and may lead to Post-COVID syndrome (PCS) symptoms.
- Patients with PCS show elevated inflammation and endothelial dysfunction markers.
- Spike proteins can persist for up to 12 months post-infection, driving ongoing inflammation and immune activation.

*What New Information Does This Article Contribute?:*

- Low haemoglobin (Hb) and high VEGF correlate with higher Anti-S1 IgG and low Hb is associated with higher C19-YRS severity score.
- PCS patients exhibit higher Erythropoietin (EPO) levels when compared to HC.
- Spike protein 1 (S1) alone and PCS patient’s plasma induce endothelial dysfunction primarily through HIF-2α activation.
- Both S1 and PCS plasma cause oxidative stress and disrupting endothelial integrity.
- Inhibition of HIF-2α effectively restores endothelial barrier integrity disrupted by S1 and PCS plasma. Persistent circulation of spike proteins can sustain chronic inflammation and immune activation in patients with PCS. Here we show that plasma from PCS patients exhibits significantly elevated levels of VEGF which positively correlates with Anti-S1 IgG. Low haemoglobin was associated with higher Anti-S1 IgG titres and correlated with a higher C19-YRS severity score. Levels of EPO were higher in PCS patients, with a more pronounced effect observed in patients with cardiovascular symptoms. In human retinal endothelial cells, both S1 and plasma from PCS patients primarily induce ED through HIF-2α activation, rather than NF-κB. Both factors lead to significant oxidative stress, evidenced by increased ROS production which in turn disrupts endothelial barrier integrity and function. Notably, Belzutifan, a HIF-2α inhibitor, can restore this compromised endothelial function, offering a potential therapeutic target for PCS.

**Graphical Abstract:** 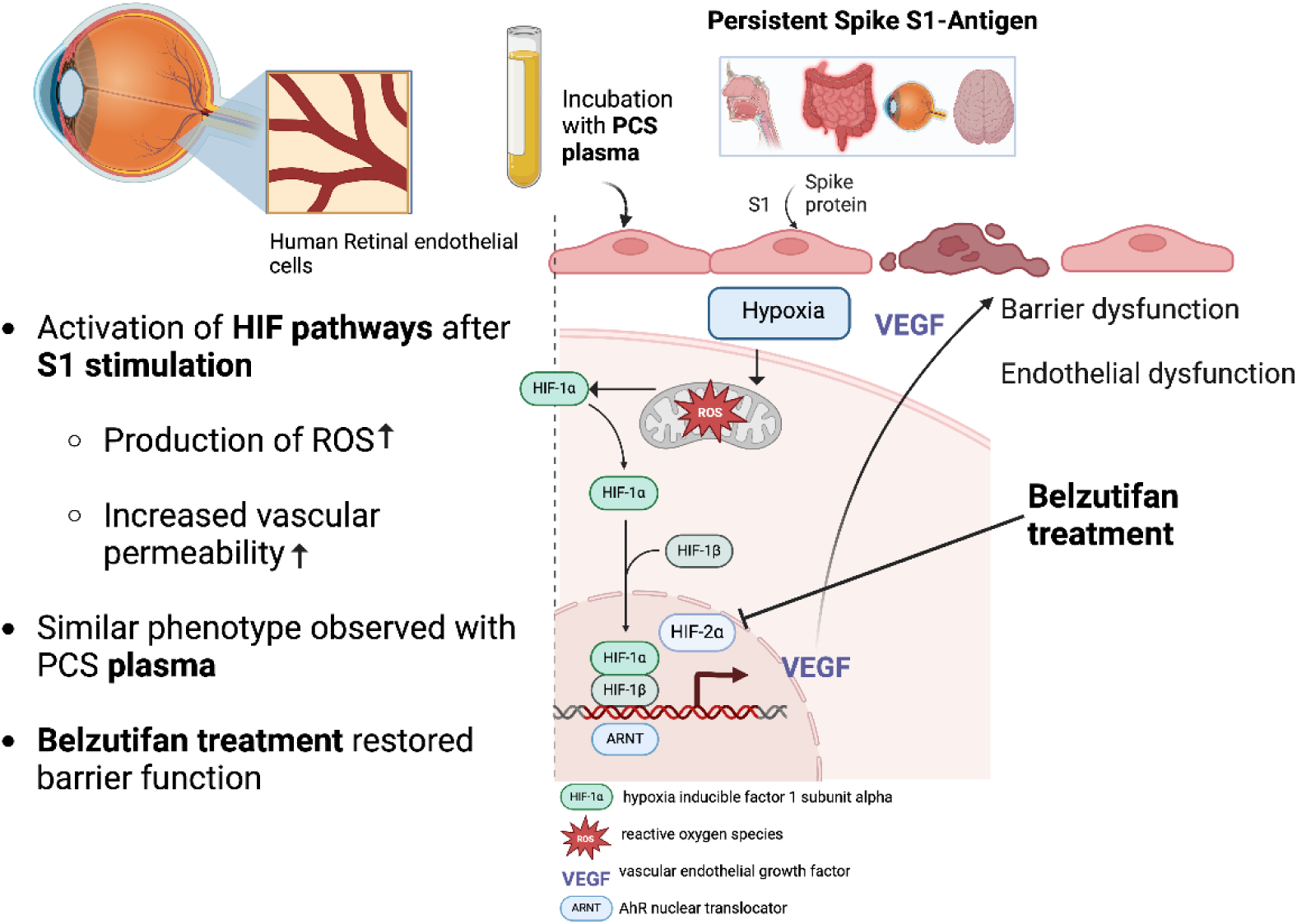

## Introduction

Some patients experience lingering symptoms known as Post-COVID (PCS), affecting 10-30% of individuals after acute SARS-CoV-2 infection [1–3]. The exact causes of PCS are still uncertain, with hypotheses including virus or viral remnants persistence [4, 5], autoimmunity [5–8], microbiota impacts [5, 9], reactivation of dormant infections unrelated to SARS-CoV-2 [10, 11], chronic inflammation-induced tissue damage [12], and ongoing cardiovascular effects potentially driven by microthrombi and persistent vascular dysfunction [13, 14].

In acute SARS-CoV-2 infection, endothelial dysfunction (ED) is common and may contribute to the persistence of symptoms in PCS [15–17]. SARS-CoV-2 can damage endothelial cells directly through the angiotensin-converting enzyme-2 (ACE2) receptor or indirectly through cytokine storm, leading to oxidative stress, reduced nitric oxide (NO) availability, and endothelial barrier disruption [18–21]. Evidence suggest that viral replication can persist for months in various tissues, potentially serving as reservoirs for spike proteins [22–24]. Spike proteins may persist in the body for up to 12 months, sustaining chronic inflammation and immune activation [4, 25]. Hypoxaemia is a common complication of acute SARS-CoV-2 infection. In the acute phase the activation of hypoxia-inducible factor-1α (HIF-1α) through reactive oxygen production (ROS) plays an important role in aggravating immune response and inflammation [26]. The two major HIFs, HIF-1 and HIF-2 are transcription factors that activate the adaptive hypoxic response and influence endothelial function by triggering the expression of vascular endothelial growth factor (VEGF) [27–29]. Interestingly also in PCS patients ongoing brain hypoxemia has been linked to neurocognitive impairment and hyperbaric oxygen therapy has been proposed to improve cognitive function [30, 31]. The recently approved HIF-2α inhibitor Belzutifan (MK-6482, previously known as PT2977) shows promise for restoring endothelial function in HIF2α-driven pathologies due to its good tolerance and low degree of toxicity [32, 33]. In this study we hypothesized that circulating S1 antigen in PCS patients is involved in the pathophysiology of ED, potentially by the activation of hypoxic response. Plasma from PCS patients with high titres of anti-S1 antibodies was able to trigger increased production of ROS, reduce NO bioavailability and activate HIF pathways. These changes could compromise endothelial barrier function and potentially promote angiogenesis. Furthermore, we aimed to investigate whether inhibiting the HIF pathway with Belzutifan can restore ED in PCS patients.

## Methods

### Clinical Assessment

This study is part of the “All Eyes on PCS” study which is an observational, single-center study studying the underlining causes of PCS. The study protocol with a detailed description of the methods has previously been published [34]. It has been written following the Strengthening the Reporting of Observational studies in Epidemiology (STROBE) guidelines [35]. A standardized questionnaire on the day of recruitment, detailed clinical evaluations and patient-reported outcome measures, were used to assess the PCS severity score [36] and the COVID-19 Yorkshire Rehabilitation Scale (C19-YRS) [37]. The study was performed in accordance with the Declaration of Helsinki and approved by the local ethics committee (Ethics Committee of the Technical University of Munich, School of Medicine, Klinikum rechts der Isar; Approval number: 2022-317-S-SR) and was previously registered (https://clinicaltrials.gov/ct2/show/NCT05635552). All participants in this study provided written informed consent.

### Laboratory values

Blood sampling was performed as previously described [38]. Standard laboratory measurements were performed in an ISO-certified routine laboratory. VEGF, IL-6, ICAM-1, VCAM-1, MCP-1, XCL10, and RANTES in the patient’s plasma were quantified using the Cytometric Bead Array Flex system (BD Biosciences, San Diego, US), according to the instructions of the manufacturer. Samples were acquired on FACSCanto II flow cytometer and sample data analysis was accomplished using FCAP array software. To determine SARS-CoV-2 Spike RBD IgG levels in plasma the enzyme-limited immunosorbent assay (ELISA) method was performed using a Legend Max ELISA kit (BioLegend) following the manufacturer’s recommendations. Erythropoietin (EPO) measurements were assessed using a Quantitative IVD ELISA (R&D systems) under the manufacturer’s instructions. Absorbances were measured at 450 nm in a microplate reader Multiskan FC (Thermo Scientific).

### Cell culture

Primary human retinal microvascular endothelial cells (HREC) were purchased from Innoprot. Cells were cultured in endothelial basal medium supplied with 5% of fetal bovine serum, 1% of endothelial cell growth supplement and 1% penicillin/streptomycin solution (all Innoprot). Cells were cultured and used from passage 2 to 6 and then discarded. Cells were maintained in a humidified incubator at 37 °C and 5% CO_2_. For experimental procedures cells were treated with medium control or active recombinant human coronavirus SARS-CoV-2 Spike glycoprotein S1 (S1) (Abcam) in the presence or absence of Belzutifan (MedchemExpress) at 50 nM. We used S1 subunit concentration of 100 ng/mL based on a previous study that tested the effects of S1 on stimulating human endothelial cells, which went from a concentration similar to that found in the sera of severe COVID-19 patients to higher concentrations used in previous studies [39]. Hypoxic conditions were achieved using CoCl2 at 100 µM as described [40]. When cells were stimulated with LPS (Invivogen), it was used at a concentration of 100 ng/mL. For cells exposed to plasma from Post-COVID patients or healthy individuals, 2% of plasma was added to medium with 3 IU/mL of heparin.

### RNA extraction, reverse transcription, and quantitative real-time PCR

Total RNA was extracted from HREC using the Norgene Total RNA Purification kit (Norgen Biotek), following the manufacturer’s protocol. The cDNA was synthesized from 1µg of total RNA by reverse transcription polymerase chain reaction (PCR) using Superscript II reverse transcriptase (Thermo Fisher) as per the manufacturer’s guidelines. The qRT-PCR from cDNA was performed in a Light Cycler 480 (Roche). GAPDH mRNA was used as a reference transcript for relative quantification. The 2ΔCT method was used to assess the expression levels of target genes in each sample. ΔCT values were calculated by subtracting the CT value of the target gene from that of the reference gene. The 2ΔCT values were then derived by raising 2 to the power of the ΔCT value, providing the relative expression levels of the target gene in each experimental sample. Fold induction was not applied for comparisons of gene expression within samples. The resulting 2ΔCT values were then subjected to statistical analysis to identify significant differences in gene expression between groups. Controls consisting of ddH2O were negative for targets and reference genes. The melting curve profiles were analyzed. All primers (Supplemental Table 1) used for amplification were purchased from Metabion.

### Immunofluorescence experiments

HREC were grown in an 8 well chamber slide previously coated with fibronectin (Innoprot) 2 µg/cm^2^. Once HREC formed a confluent monolayer, cells were submitted to the desired treatment. After treatment, monolayers of HREC were fixed in 4% formaldehyde for 15 minutes at room temperature. Cells were then treated with permeabilization buffer containing 0,1% TritonX-100 in PBS for 10 minutes at room temperature followed by the treatment with blocking buffer containing 3% BSA and 0,1% TritonX-100 in PBS for 30 minutes at room temperature. Subsequently, cells were incubated with the primary antibodies for NF-kB, HIF-1α, HIF-2α, BNIP-3, GLUT-1, F-actin, Claudin-5, VE-Cadherin (Cell Signaling) or PDK-1 (Abcam), in an antibody dilution buffer containing 1% BSA and 0,1% TritonX-100 in PBS, overnight, at 4 °C. Next, cells were washed three times in PBS and incubated with a fluorescently-tagged goat anti-rabbit secondary antibody (Cell Signaling), in antibody dilution buffer containing 1% BSA in PBS for 1 hour at room temperature protected from light. Cells were then washed three times in PBS and incubated with phalloidin (Abcam) diluted in 1% BSA in PBS for 90 minutes. The cells were subsequently washed three times with PBS and DAPI (Vector Laboratories) was added. Images were acquired in a Leica DM RBE fluorescence microscope. Corrected Total Cell Fluorescence (CTCF) was calculated for each cell by applying the background-subtracted integrated density values using the formula: CTCF = Integrated Density - (Area × Mean Fluorescence of Background). All measurements, including the integrated density, cell area, and background fluorescence, were performed using ImageJ. Measurements were taken from multiple cells within each condition to ensure statistical reliability. Background fluorescence was determined by selecting regions without cells in each image as background regions of interest.

### Measurement of ROS

Cellular ROS generation was assessed using DCFDA/H2DCFDA-Cellular ROS assay kit (Abcam). ROS in the cells cause oxidation of DCFH, yielding the fluorescent product 2′,7′-dichlorofluorescein (DCF). HREC were seeded in a dark clear-bottom 96-well microplate (Corning) at a density of 25,000 cells per well and allowed to adhere overnight. Subsequently, cells were incubated with DCFDA for 45 minutes in the dark. Following DCFDA removal, cells were exposed to the desired stimuli, and fluorescence intensity was measured at Ex/Em= 485/535 nm at 60 minutes intervals over a period of 6 hours. Mitochondrial ROS generation was evaluated using MitoSOX Red Mitochondrial indicator (Invitrogen) as described [41]. HREC were exposed to the intended treatment for a period of 4 hours. Following treatment, the cells were harvested into flow cytometry tubes and subjected to incubation with MitoSOX red reagent for 20 minutes at 37 °C in a light-protected environment. Following the incubation period, the cells were prepared for flow cytometry analysis, with the quantification of MitoSOX expression being measured in the PE channel.

### Measurement of NO

Total NO production in supernatants was assessed using a commercially available fluorometric Nitric Oxide assay kit (Sigma-Aldrich) and according to the manufacturer instructions. The assay is based on the enzymatic conversion of nitrate to nitrite by nitrate reductase, followed by the addition of 2,3-diaminonapthalene (DAN) and NaOH, which converts nitrite to a fluorescent compound. Briefly, 100 µLof supernatant from cells exposed to medium control, S1, HC-plasma or PCS-plasma, diluted in assay buffer, were dispensed into a 96-well plate. 10 µL of enzyme co-factors and 10 µL of nitrate reductase were added to each well. The plate was incubated at room temperature for 1 hour. After the required incubation period, 10 µL of DAN reagent was added to each well and the plate was incubated for 10 minutes followed by the addition of 20 µL of NaOH to each well. Fluorescence intensity was measured at Ex/Em= 375/415 nm. The nitrate+nitrite concentration was caPCSulated from a nitrate standard curve.

### Transendothelial electric resistance (TEER) measurement

The barrier function of confluent endothelial cell monolayers was estimated using electric cell-substrate impedance sensing (ECIS) model Z-Theta (Applied Biophysics) as described [42]. HREC monolayers were grown on standard 8-well array (8W10E + PET) (Ibidi). Resistance was measured continuously in a multi-frequency setup. After the resistance at 4,000 Hz reached a stable plateau of >1,000-ohm, HREC were incubated with fresh medium in the presence or absence of S1 or Belzutifan and the capacity of the barrier integrity disruption was measured over a 72 hours period. On a second approach, plasma samples from HC or LC patients were diluted to 2% (v/v) in cell culture medium in the presence or absence of Belzutifan and tested for a period of 42 hours. After treatment, cells were continuously monitored in real-time at multiple frequency modes. Values were normalized to time = 0 for easier comparisons.

### Statistical analysis

Statistical calculations were performed using GraphPad Prism 10 software and R Studio (Version 2024.04.2+764). The graphical abstract was generated using BioRender.com. Further details on statistical analysis are provided in the Supplemental Material

## Results

### Main baseline characteristics of PCS cohort

The study included 41 Post-COVID Syndrome (PCS) patients (mean age 42.2 years, ±12.2, 75.6% female) who had previously been infected with SARS-CoV-2 and exhibited PCS typical symptoms. Patients were age- and gender-matched with 24 healthy volunteers recruited before the COVID-19 pandemic (mean age 44.6 years, ±12.2, 70.8% female; HC group) (Figure 1A). The prevalence of cardiovascular risks, such as arterial hypertension, nicotine abuse, diabetes mellitus and obesity was similar between the groups. 73.2% of the PCS patients had one infection and 26.8% had two infections with Omicron (24.4%) being the most frequent variance. 23 (56,1%) patients were vaccinated three times, 15 (36.6%) patients were vaccinated twice, and 3 (7.3%) patients were not vaccinated. Anti-S1 IgG levels did not vary with the number of infections (Figure 1B) or vaccinations (Figure 1D). The median duration of PCS was 12.7 months (±8.4). Among the PCS patients, 19.5% experienced work loss, with a median sick leave duration of 122.0 days (4.0 - 291.0 days). Most common symptoms reported by PCS patients included fatigue (95.1%), exercise intolerance (90.2%) and brain fog (90.2%) (Figure 1C). Eight PCS symptom clusters were identified (Figure 1E) and the mean severity score measured by C19-YRS was 38.20 (±18.21). Anti-S1 IgG levels were significantly higher in PCS patients compared to HC (Supplemental Table 2). EPO levels were significantly higher in PCS patients compared to HC (Figure 1F). Additionally, PCS patients with two or more cardiovascular (CV) symptoms (Palpitations, shortness of breath, reduced physical resilience and chest pain) had significantly higher EPO levels than those with less than two CV symptoms (Figure 1G). To address a potential link between dysregulated erythropoiesis, S1 levels and PCS severity we correlated Hb and haematocrit (Hct) with Anti-S1 IgG levels and C19-YRS. Both Hb and Hct showed a non-significant negative correlation with Anti-S1 (Supplemental Figure 1B and C). After controlling for confounders low levels of Hb were significantly associated with high levels of Anti-S1 (Supplemental Table 3). Both, lower levels of Hb (Supplemental Figure 1D) and Hct (Supplemental Figure 1E) were significantly correlated with a higher C19-YRS severity score. The association remained significant for Hct, however was weaker for Hb after controlling for potential confounders (Supplemental Table 3).

**Figure 1.**
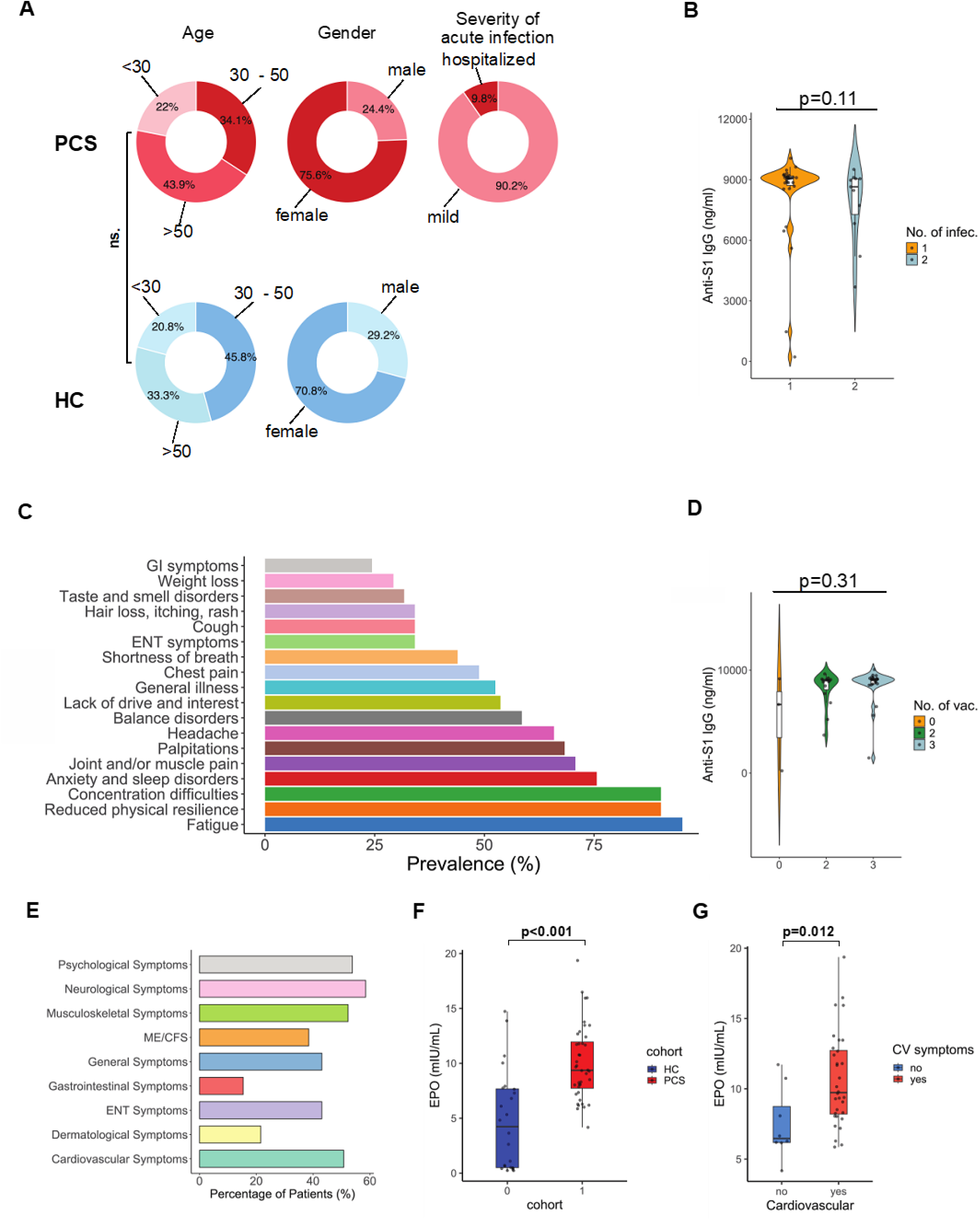
Visual presentation of demographics and Post-COVID Syndrome symptoms in cohort. **(A)** Pie chart shows distribution of age and gender in the Post-COVID (PCS) cohort (n=41, top, red color scale) and healthy cohort (HC) (n=24, bottom, blue color scale) and the distribution of the severity of the acute infection in the PCS cohort. Statistical testing was done using the χ2 test for categorical values and the Students t-test was used for parametric variables. **(B)** Violin plots show Anti-S1 IgG for Number (No.) of infections (inf.). Testing was done using the Mann-Whitney t test for non-parametric distribution. **(C)** Prevalence of the top 18 self-reported PCS symptoms ranked from most prevalent (bottom) to least prevalent (top). **(D)** Violin plots show Anti-S1 IgG for Number (No.) of vaccinations (vac.) in PCS cohort. Testing was done using the Kruskal Wallis test for non-parametric distribution. **(E)** Bars show 8 PCS symptom clusters: Psychological (Lack of interest and anxiety and sleep disorders), gastrointestinal (Abdominal pain, diarrhoea, vomiting, nausea), dermatological (Hair loss, itching, rash), neurological (headache, concentration difficulties), cardiovascular (palpitations, chest pain, shortness of breath, reduced physical resilience), ear-nose-throat (ENT; cough, taste and smell disorders, sore throat, runny nose, hoarseness), musculoskeletal (joint and/or muscle pain, balance disorders) and general (fatigue with a Fatigue severity socre >5, chills/fever, flu-like feeling, weight loss) symptoms. **(F and G)** Box plots compare Erythropoietin (EPO) levels in PCS patients versus HC and in PCS patients with two or more cardiovascular (CV) symptoms versus PCS patients with less than two CV symptoms. Testing was done using the Mann-Whitney t test for non-parametric distribution.

### Elevated plasma levels of ED and inflammation markers in PCS

In a prior study we demonstrated that prolonged ED is a hallmark of PCS, potentially linked to chronic inflammation [17]. We profiled the plasma samples from the participants for a panel of seven markers including VEGF, IL-6, ICAM-1, VCAM-1, MCP-1, CXCL10, and RANTES. Of the selected markers, VEGF and MCP-1 were significantly higher in PCS patients (Figure 2A and 2C). RANTES, ICAM-1, VCAM-1, and CXCL10 did not show a significant increase in PCS-plasma when compared with HC-plasma (Figure 2D to 2G). Surprisingly, IL-6 showed lower levels in plasma samples from PCS patients when compared with the HC. However, with both groups showing IL6 values in the normal range. (Figure 2B).

**Figure 2.**
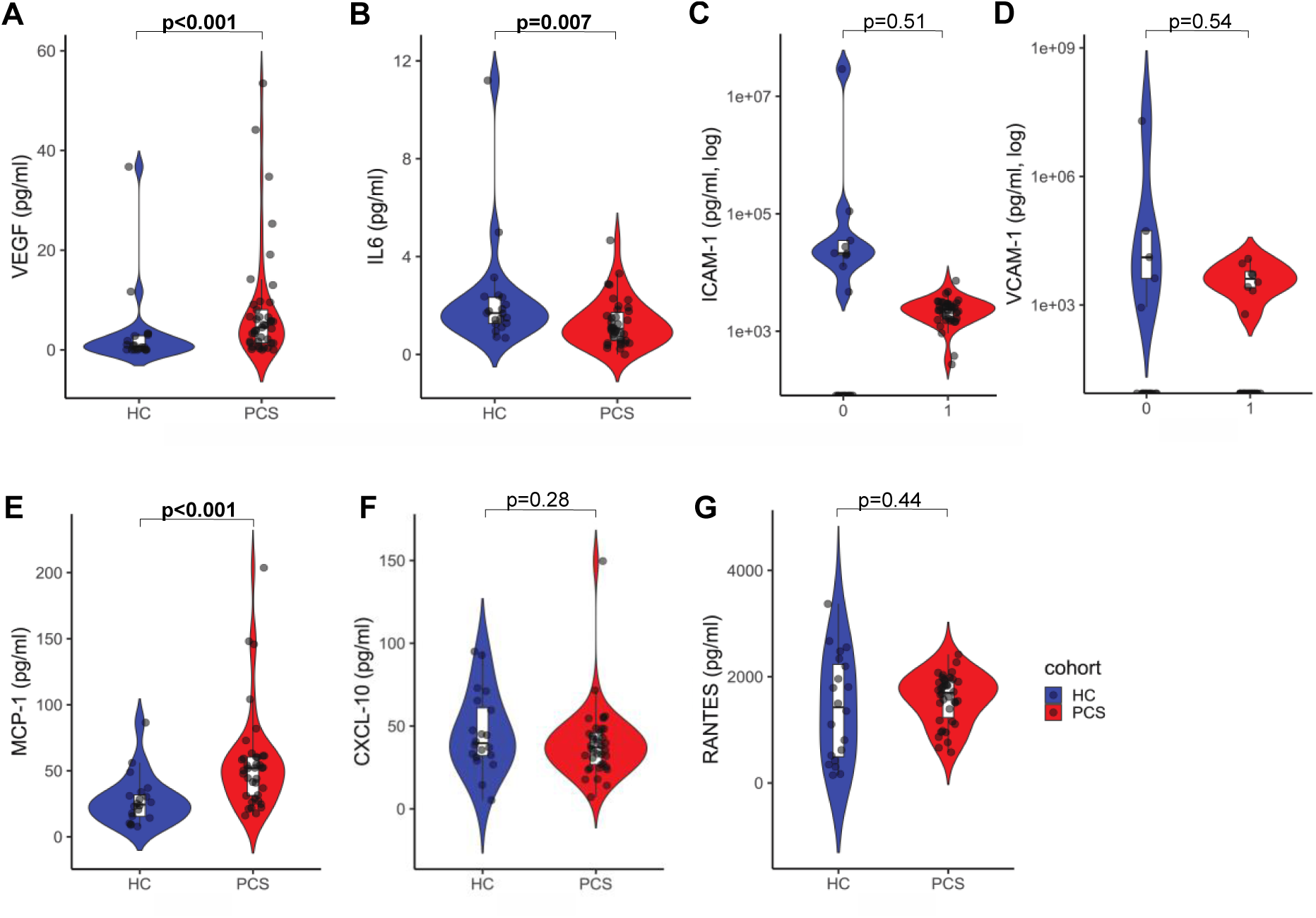
Parameters of inflammation and ED in patients with PCS compared to HC. Violin plots comparing **(A)** VEGF, **(B)** IL-6, **(C)** ICAM-1, **(D)** VCAM-1, **(E)** MCP-1, **(F)** CXCL-10, and **(G)** RANTES levels between PCS patients and the healthy cohort (HC). Plots show values with the mean indicated by a line. VEGF was measured in a total of n=41 PCS patients and n=20 HC; IL-6, MCP-1, RANTES and CXCL-10 in a total of n=38 in PCS patients and n= 20 HC. The Mann-Whitney *t* test was used for non-parametric distributions.

Given reports of higher Anti-S1 IgG in participants with PCS, we correlated VEGF and inflammatory markers with Anti-S1 IgG levels. Inflammatory markers such as IL6, RANTES, MCP-1 and CXCL10 did not show a significant correlation (not shown). Higher VEGF was positively correlated with Anti-S1 IgG levels (Supplemental Figure 1A), however this effect was weaker after controlling for potential confounders in a multivariate linear model (Supplemental Table 3).

### S1 induces increased VEGF mRNA expression without upregulating immune activation markers in HREC

With studies reporting persistent circulation of spike protein in PCS patients, we studied the overall effect of SARS-CoV-2 recombinant S1 spike protein (S1) on the gene expression of markers indicating ED and immune activation in primary human retinal endothelial cells (HREC). Therefore, HREC were exposed to either medium (negative control), S1 and LPS (positive control) for 4 hours, and the mRNA expression of VEGF, IL-6, TNF, IL-8, MCP-1, CXCL1, ICAM-1, and CXCL10 was assessed by quantitative PCR. Notably, exposure to S1 did not induce significant upregulation in IL-6, TNF, MCP-1, CXCL1, ICAM-1 and CXCL-10 mRNA expression (Figure 3B and 3C, Figure 3E to 3H). However, VEGF exhibited a substantial mRNA expression following treatment with S1 (Figure 3A). In contrast, IL-8 showed a significant decrease after S1 exposure (Figure 3D).

**Figure 3.**
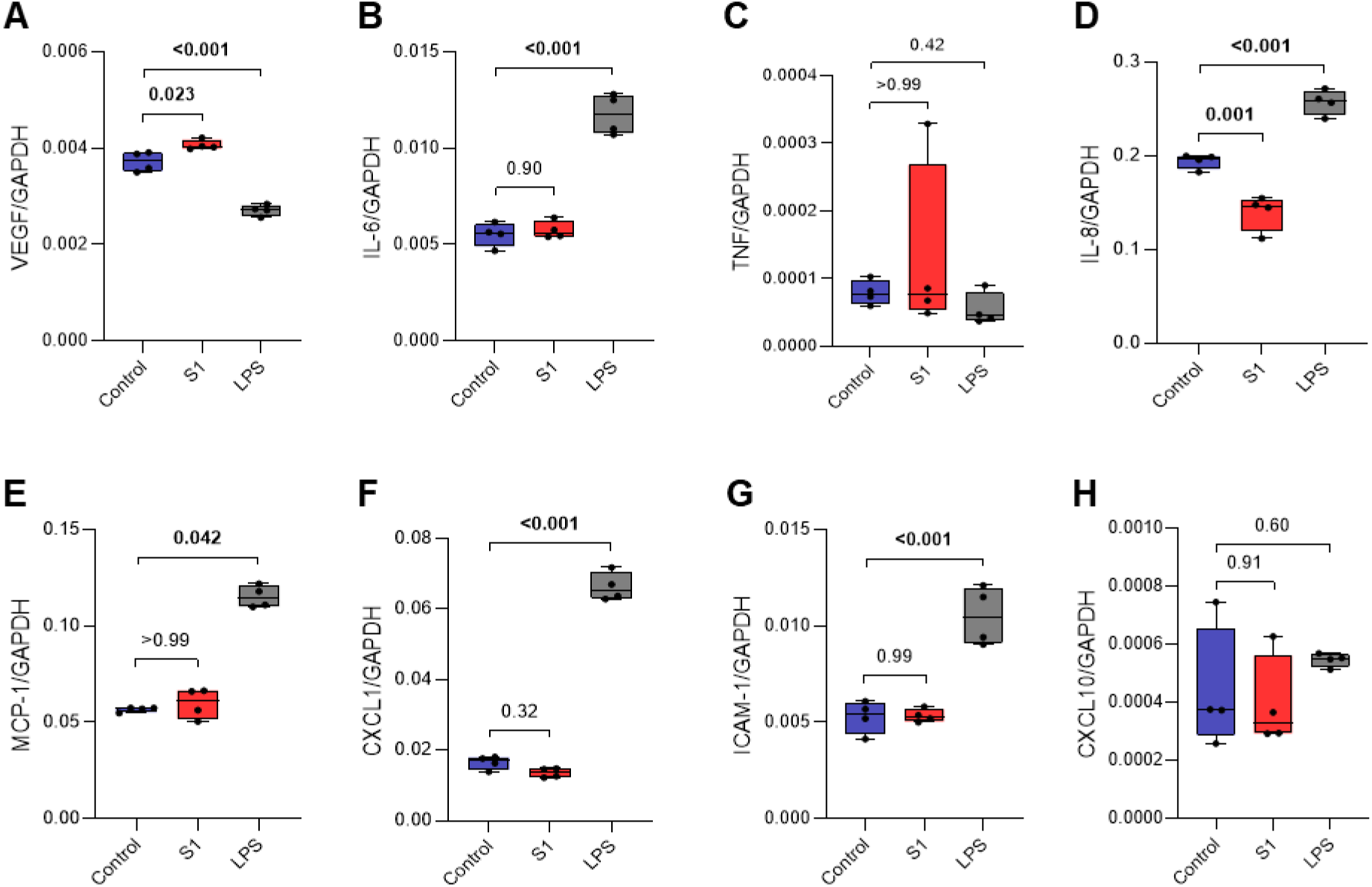
Effect of recombinant SARS-CoV-2 S1 spike protein on gene expression of ED and inflammatory markers in HREC. Quantitative real-time PCR was performed to determine the mRNA expression of a set of inflammatory markers in HREC. Data are shown as mean ± SD of the expression values normalized to the GAPDH control for **(A)** VEGF, **(B)** IL-6, **(c)** TNF, **(D)** IL-8, **(E)** MCP-1, **(F)** CXCL1, **(G)** ICAM-1, and **(H)** CXCL10 mRNA in HREC exposed to medium (control, n=4), 100 ng/mL of S1 (n=4) and 100 ng/mL LPS (n=4) for 4 hours. A p-value of <0.05 was considered statistically significant. P values were determined by 1-way ANOVA followed by Tukey’s post hoc test **(A, B, G, F G and H)** or Kruskal-Wallis test followed by Dunn’s post hoc test **(C and E).**

### S1 leads to HIF-1α and HIF-2α nuclear activation and downstream pathways rather than NF-κB in HREC

Given the observation that S1 fails to elicit the selected inflammatory markers in HREC but VEGF exhibits substantial upregulation, we assessed the activation status of nuclear factor-kappa B (NF-κB) and HIF-1/2α in cells treated with S1. LPS, a robust activator of the NF-kB signalling pathway, and cobalt chloride (CoCl2), a potent inducer of hypoxia, were used as positive-controls, respectively. Nuclear translocation of NF-kB was evident at 4 hours post-treatment with LPS (Supplemental Figure 2). In contrast, cells treated with S1 exhibited a markedly less pronounced response without significance compared to the control (Supplemental Figure 2) confirming our previous results that S1 has a limited impact on the expression of the selected inflammatory markers. Nuclear expression of HIF-1α was absent in cells treated with the medium control. Cells treated with either recombinant S1 or CoCl2 exhibited a significant activation of HIF-1α at 8 hours after treatment (Figure 4A).

**Figure 4.**
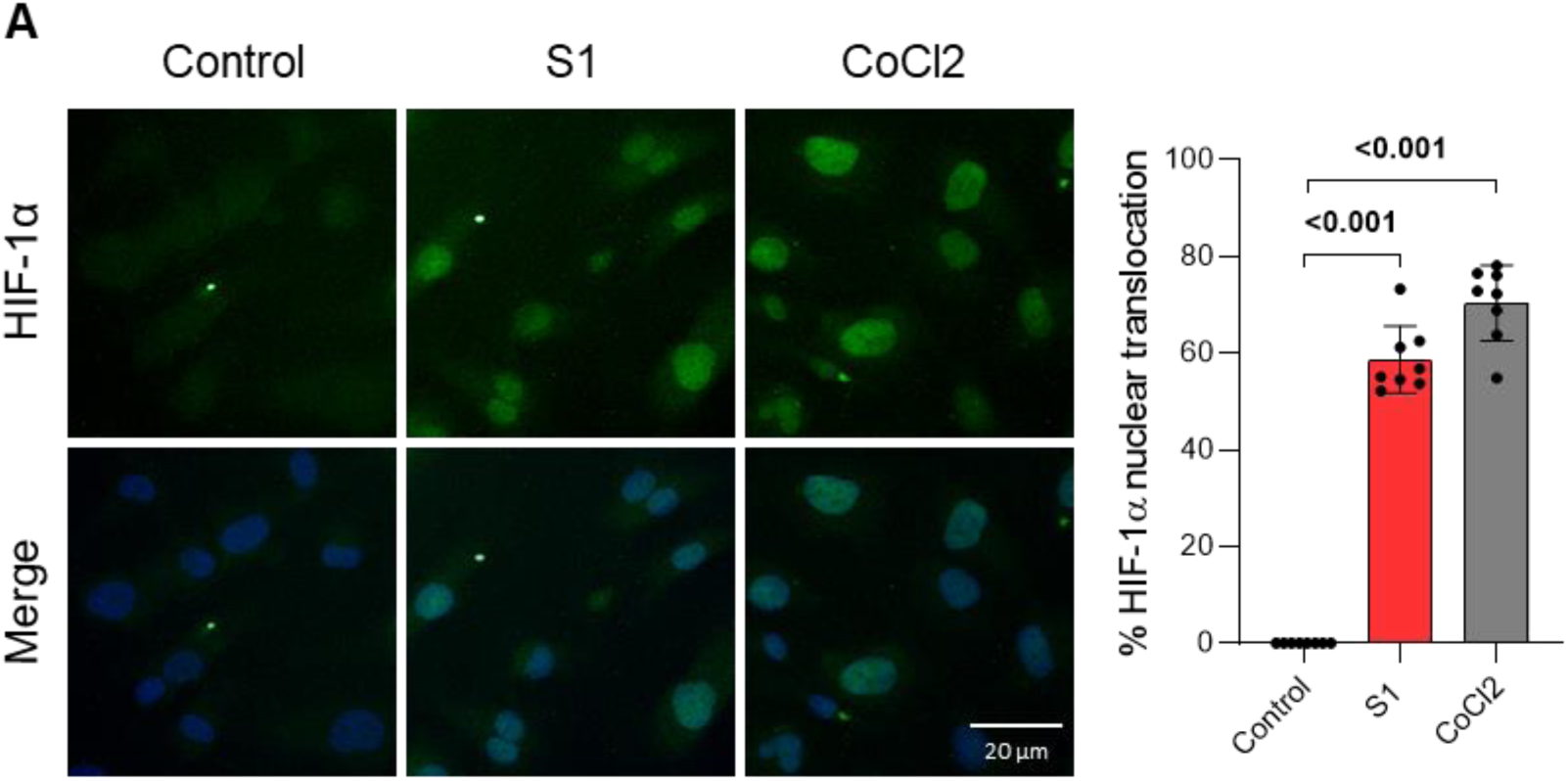
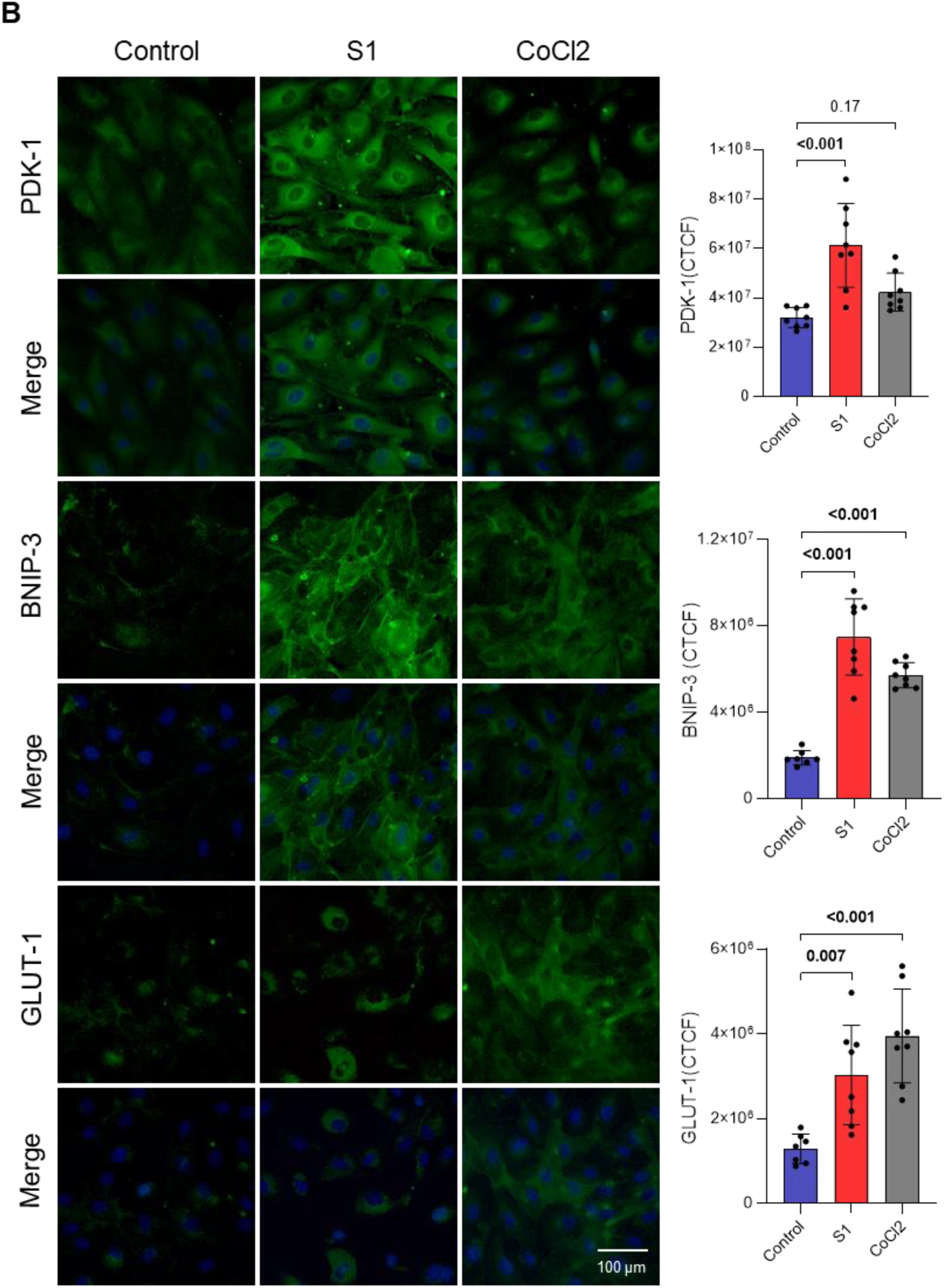
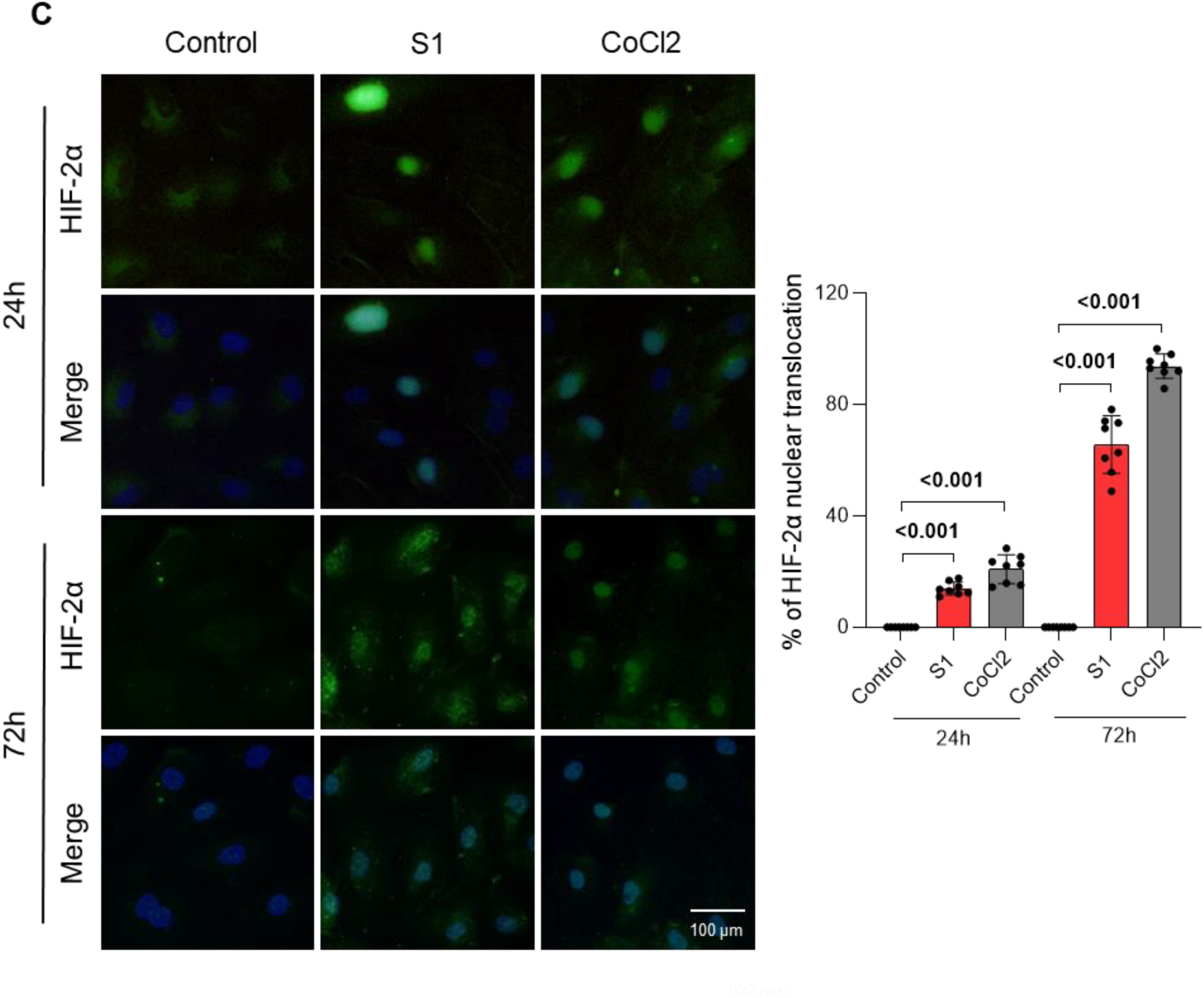
S1 induces high HIF-1/2α nuclear translocation in HREC. Representative confocal analysis of **(A)** HIF-1α in HREC treated with medium (Control, n=8), 100 ng/mL S1 (n=8), and 100 µM CoCl2 (n=8) for 8 hours. **(B)** PDK-1, BNIP-3, and GLUT-1 in HREC exposed to medium (Control, n=7), 100 ng/mL S1 (n=8), and 100 µM CoCl2 (n=8) for 24 hours; **(C)** HIF-2α in HREC treated with medium (Control, n=8), 100 ng/mL of S1 (n=8), and 100 µM CoCl2 (n=8) for 24 hours or 72 hours. All the primary antibodies were labelled with FITC (green) while the nuclei were counterstained with DAPI (blue). Images (left) were acquired at 20× magnification and scale bars represent 20 µm **(A and C)** or 100 µm **(B)**. Graphs (right) illustrate the percentage of nuclear translocation **(A and C)** and the corrected total cell fluorescence **(B).** Data are shown as means ± SD. A p-value of <0.05 was considered statistically significant. P values were determined by 1-way ANOVA followed by Tukey’s post hoc test.

To investigate the activation of HIF transcriptional pathways, we analysed the expression of key HIF target proteins including phosphoinositide-dependent protein kinase 1 (PDK1), BCL2/adenovirus E1B 19 kDa protein-interacting protein 3 (BNIP3) and Glucose transporter 1 (GLUT1) by immunofluorescence. Our findings revealed that HREC express high levels of the three selected markers under S1 exposure (Figure 4B).

S1 induced HIF-1α expression is dramatically reduced at 24 hours post-treatment (Figure 4A and Supplemental Figure 3). In contrast HIF-2α is activated after 24 hours exposure to S1 an effect that is exacerbated after 72 hours (Figure 4C). This observation underscores the capacity of S1 to induce HIF-1/2α nuclear expression.

### S1 induces ROS production, affects NO availability and disrupts retinal endothelial monolayer integrity, an effect restored by Belzutifan

Next we tested whether S1 exposure affects ROS and NO production in HREC. ROS significantly increased in a time-dependent manner (3 hours, p=0.014; 4 hours, p= 0.008; 5 hours, p=0.010; 6 hours, p=0.025) upon S1 treatment (Figure 5A). Additionally, mitochondrial ROS production also rose in cells treated with S1 (Figure 5B). S1 reduced NO production, which however failed to reach significance. (Supplemental Figure 4).

**Figure 5.**
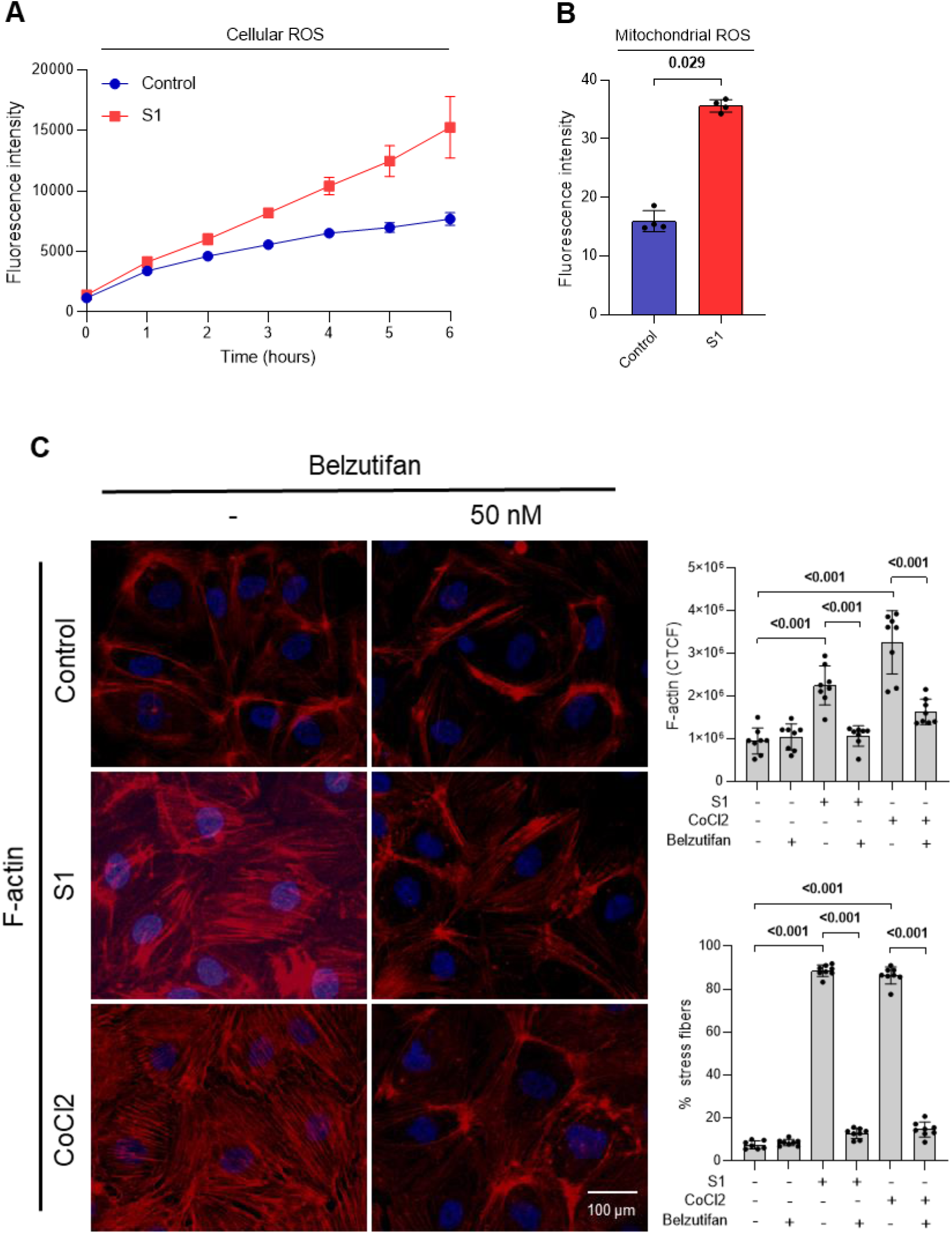
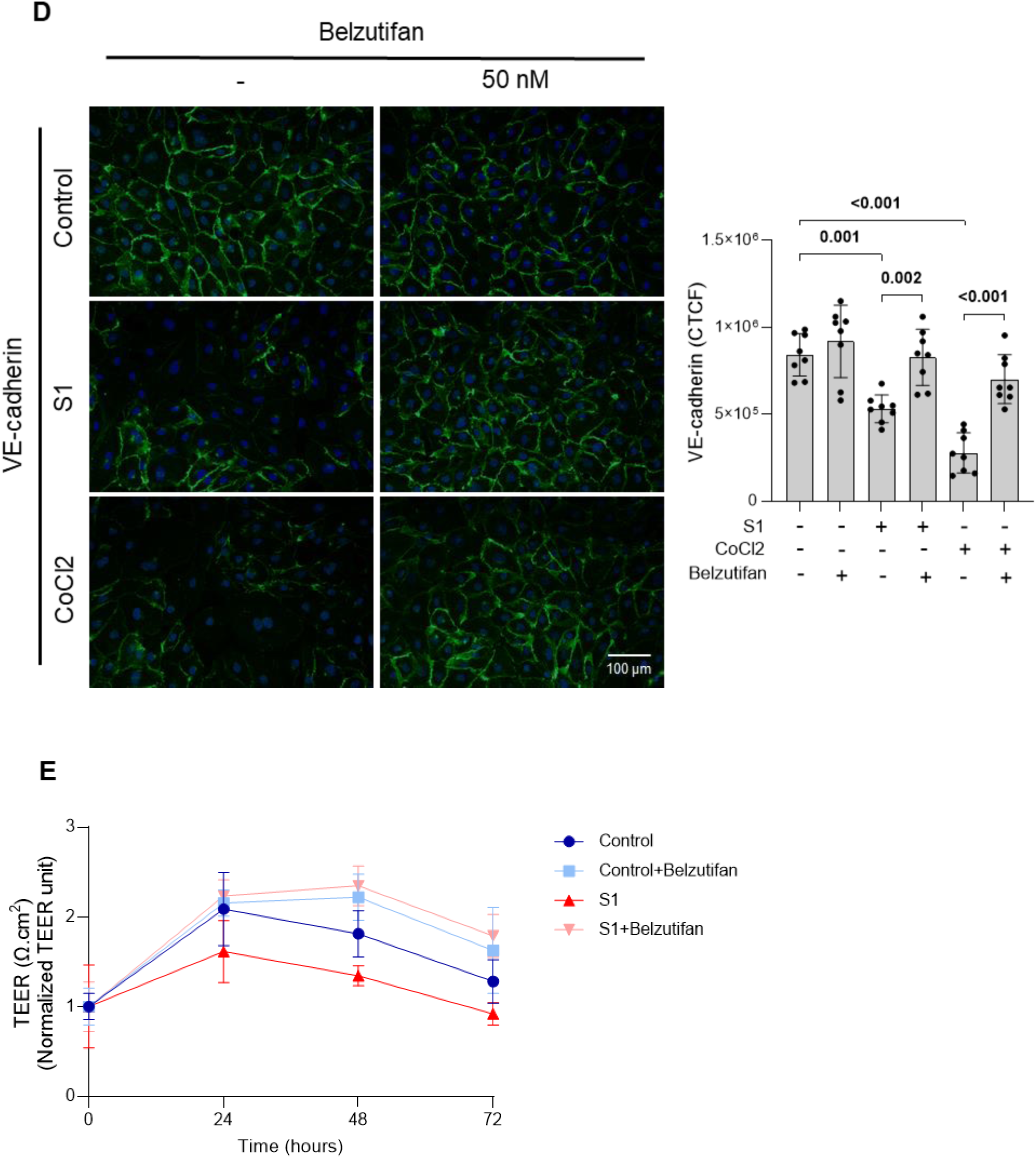
Effects of S1 on ROS production and barrier integrity in HREC and the restorative effect of Belzutifan. **(A)** Measurement of cellular ROS levels in HREC treated with medium (Control, n=3) and S1 at 100 µg/mL (n=3) in a time dependent treatment, from 0 to 6 hours, using DCFDA/H2DCFDA. **(B)** Quantitative analysis of MitoSox red fluorescence in HREC exposed to medium (Control, n=4) and 100 µg/mL S1 (n=4) for 4 hours. **(C and D)** Immunofluorescence staining for F-actin (red) **(C)**, and VE-cadherin (green) **(D),** in HREC exposed to medium (Control, n=8), 100 ng/mL S1 (n=8), and 100 µM CoCl2 (n=8), and treated with 50 nM Belzutifan (n=8) for 72 hours. Nuclei were labelled with DAPI (blue). Images (left) were acquired at 20× magnification and scale bars represent 100 µm. Graphs (right) illustrate the percentage of positivity **(C)** and the corrected total cell fluorescence (CTCF) **(C and D)**. **(E)** HREC were cultured at confluence on ECIS electrodes and then stimulated with 100 ng/ mL S1 or left untreated in the presence or absence of 50 nM Belzutifan in a time dependent treatment from 0 to 72 hours. The loss of barrier integrity was determined by transendothelial electrical resistance (TEER). Values were normalized to time = 0 for easier comparisons. Data are shown as means ± SD. A p-value of <0.05 was considered statistically significant. P values were determined by 2-way ANOVA followed by Tukey’s post hoc test **(A and E)**, Mann-Whitney’s 2-tailed t test **(B)**, and 1-way ANOVA followed by Tukey’s post hoc test **(C and D)**.

Following our observations, we investigated whether the S1 alone is capable to alter endothelial barrier integrity and function. HREC were treated with or without S1 for 72 hours before immunofluorescence staining to detect F-actin formation and Claudin-5 and VE-cadherin distribution in HREC. F-actin in control condition localized cortically (Figure 5C). In contrast, exposure to S1 diminished cortical actin and showed an elongated and narrower cell morphology (Figure 5C). S1 treatment triggered cytoskeletal remodelling, characterized by the formation of prominent actin stress fibers (Figure 5C). Furthermore, cells treated with medium control displayed almost intact VE-Cadherin (Figure 5D) and Claudin-5 (Supplemental Figure 5) bordering cells in unbroken, confluent monolayers. Upon treatment with S1, there was a significant reduction of VE-Cadherin and Claudin-5, with gaps appearing between cells (Figures 5D and Supplemental Figure 5). In line with these findings, the transendothelial electrical resistance (TEER) of the cell monolayer significantly decreased in S1-treated cells (Figure 5E, Table 1), confirming the loss of endothelial barrier integrity.

**Table 1:**
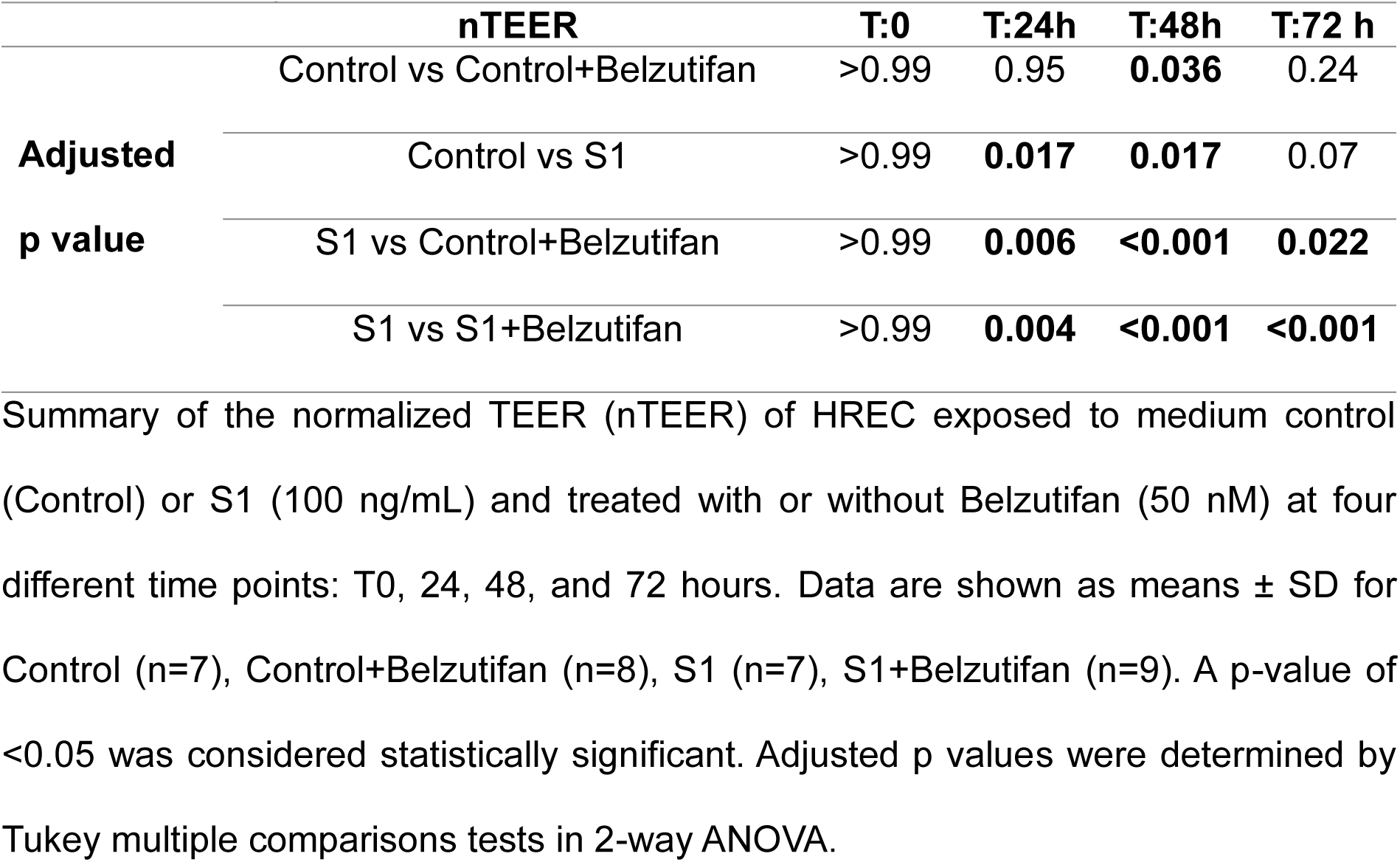
Summary of TEER in HREC exposed to S1 and treated with Belzutifan.

|  | **nTEER** | **T:0** | **T:24h** | **T:48h** | **T:72 h** |
| --- | --- | --- | --- | --- | --- |
|  | Control vs Control+Belzutifan | >0.99 | 0.95 | **0.036** | 0.24 |
| **Adjusted** | Control vs S1 | >0.99 | **0.017** | **0.017** | 0.07 |
| **p value** | S1 vs Control+Belzutifan | >0.99 | **0.006** | **<0.001** | **0.022** |
|  | S1 vs S1+Belzutifan | >0.99 | **0.004** | **<0.001** | **<0.001** |

Summary of the normalized TEER (nTEER) of HREC exposed to medium control (Control) or S1 (100 ng/mL) and treated with or without Belzutifan (50 nM) at four different time points: T0, 24, 48, and 72 hours. Data are shown as means ± SD for Control (n=7), Control+Belzutifan (n=8), S1 (n=7), S1+Belzutifan (n=9). A p-value of <0.05 was considered statistically significant. Adjusted p values were determined by Tukey multiple comparisons tests in 2-way ANOVA.

Next, we hypothesize that by inhibiting HIF-2α we could partially reverse the damage inflicted by S1 on the monolayer barrier. To validate this hypothesis, we treated HREC exposed to S1 or CoCl2 with Belzutifan for 72 hours and assessed HIF-2α activation, stress fiber production as well as VE-cadherin and Claudin-5 expression. We observed that HIF-2α activation induced by either S1 or CoCl2 was completely inhibited by Belzutifan (Supplemental Figure 6). Moreover, we observed that the excessive generation of stress fibers in cells treated with S1 or CoCl2 was significantly reduced (Figure 5C). In addition, the decline in VE-cadherin was also reversed with their levels returning nearly to those of the control (Figure 5D). Belzutifan restored Claudin-5 expression levels reduced by S1 to those similar to the control however it did not reverse the Claudin-5 reduction caused by CoCl2 (Supplemental Figure 5). Endothelial integrity of HREC was significantly higher in cells treated with Belzutifan as evidenced by TEER (Figure 5E, Table 1). The integrity of HREC exposed to S1 or Belzutifan is not induced by changes in cellular viability, as neither S1 or Belzutifan are cytotoxic or influenced the endothelial viability (Supplemental Figure 7). These findings suggest that HIF-2α is involved in S1-induced endothelial barrier impairment and dysfunction, and that Belzutifan restores monolayer integrity.

### Plasma from PCS patients increases ROS production, diminishes NO availability and induces endothelial barrier disruption in HREC, an effect reversed by Belzutifan

We used an *in vitro* approach by exposing HREC to 2% plasma from patients with PCS or HC to assess correlations between ED, increased ROS production, and impaired NO synthesis. Basic characteristics showed no significant differences between the two cohorts (Supplemental Table 4). As we observed associations between PCS severity and ED, plasma samples from PCS patients were solely selected based on their severity score. Our findings revealed a time-dependent rise in cellular ROS (1 hour, p=0.0086; 2 hours, p=0.0064; 3 hours, p=0.0.0083; 4 hours, p= 0.012; 5 hours, p=0.0321; 6 hours, p=0.0298) following treatment with PCS-plasma (Figure 6A). Additionally, mitochondrial ROS significantly increased 4 hours post-treatment (Figure 6B). Consistently, NO production decreased in HREC exposed to PCS-plasma, but only significantly after 24 hours of exposure in comparison to HC treated cells (Figure 6C).

**Figure 6.**
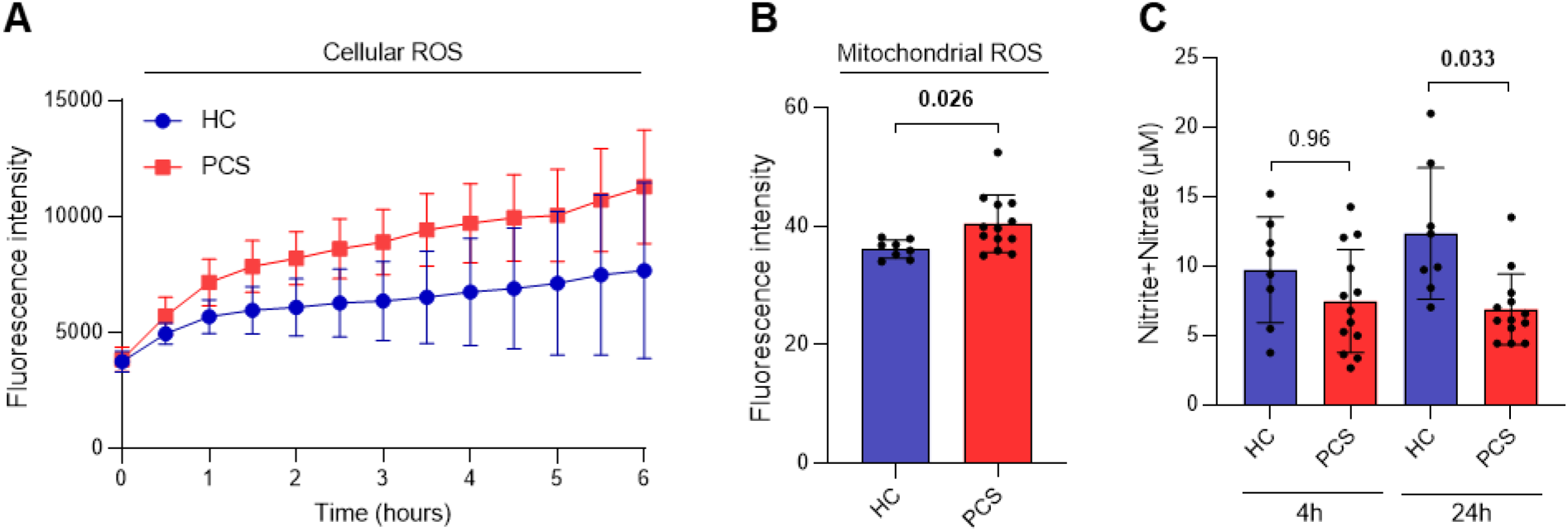
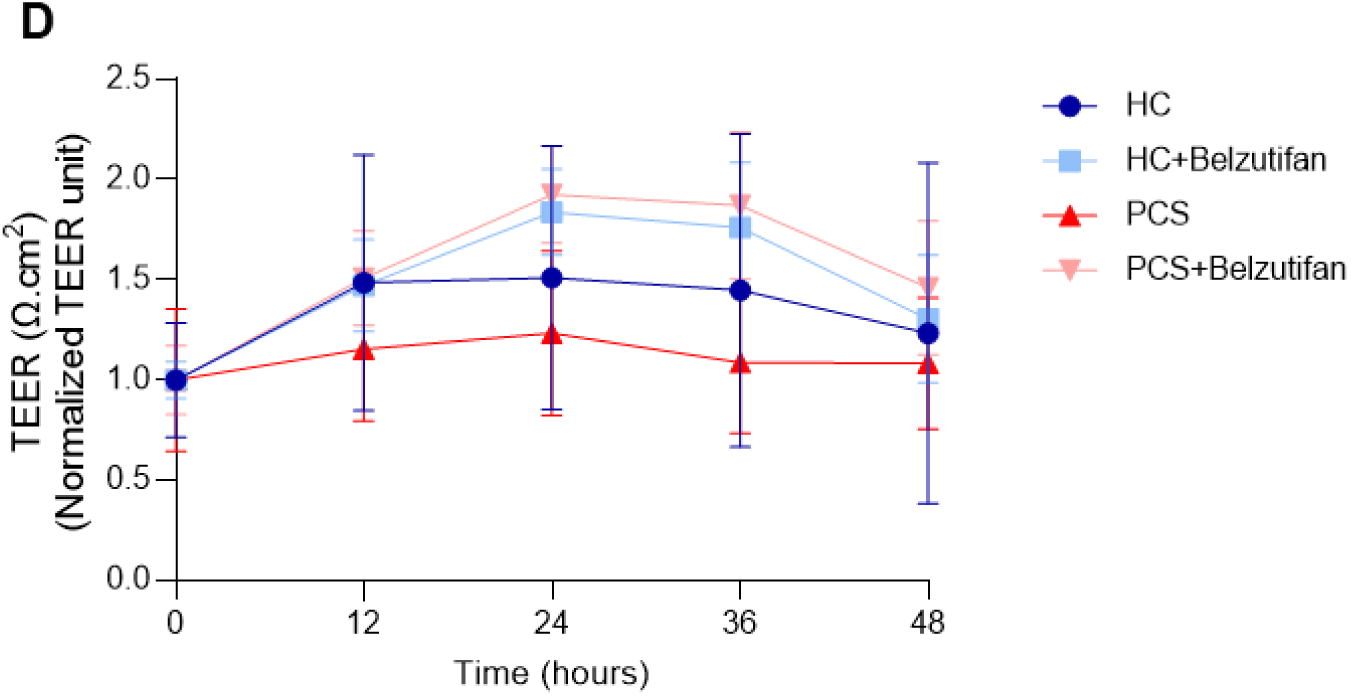
Plasma from healthy controls (HC) or Post-COVID Syndrome (PCS) patients influences ROS production, impairs NO availability and disrupts barrier integrity, an effect reversed by Belzutifan. **(A)** Measurement of cellular ROS levels in HREC treated with 2% plasma in a time dependent treatment, from 0 to 6 hours, using DCFDA/H2DCFDA. **(B)** Quantitative analysis of MitoSox red fluorescence in HREC exposed to 2% plasma for 4 hours. **(C)** Total NO level measurement of HREC exposed to 2% plasma for 4 hours, using a fluorometric-based assay that measures total nitrite/nitrate levels. **(D)** HREC were cultured at confluence on ECIS electrodes and then exposed to 2% plasma from healthy individuals (HC) or Post-COVID Syndrome (PCS) patients in the presence or absence of 50 nM Belzutifan in a time dependent treatment from 0 to 48 hours. The loss of barrier integrity was determined by transendothelial electrical resistance (TEER). Values were normalized to time = 0 for easier comparisons. Data are shown as means ± SD for n=8 healthy individuals (HC) and n=13 Post-COVID (PCS) syndrome patients. A p-value of <0.05 was considered statistically significant. P values were determined by 2-way ANOVA followed by Tukey’s post hoc test **(A and D)**, Student’s 2-tailed t test **(B)**, and Kruskal-Wallis test followed by Dunn’s post hoc test **(C).**

To investigate whether plasma from PCS patients would decrease TEER, we conducted ECIS experiment. We observed a non-significant trend towards decreased TEER in cells treated with PCS-plasma mirroring the effect observed with S1 treatment (Figure 6D, Table 2). Belzutifan again effectively restored the barrier integrity in cells treated with PCS-plasma as shown by the significant increase of TEER (Figure 6D, Table 2).

**Table 2:** Summary of TEER in HREC exposed to plasma from PCS patients and treated with Belzutifan.

|  | **nTEER** | **T:0** | **T:12 h** | **T:24h** | **T:36 h** | **T:48 h** |
| --- | --- | --- | --- | --- | --- | --- |
|  | HC vs HC+Belzutifan | >0.99 | 0.99 | 0.32 | 0.51 | 0.99 |
| **Adjusted** | HC vs PCS | >0.99 | 0.65 | 0.74 | 0.59 | 0.97 |
| **p value** | PCS vs HC+Belzutifan | >0.99 | 0.37 | 0.11 | 0.12 | 0.49 |
|  | PCS vs PCS+Belzutifan | >0.99 | **0.007** | **0.003** | **0.008** | 0.13 |

Summary of the normalized TEER (nTEER) of HREC exposed to 2% of plasma from Post-COVID Syndrome patients (PCS) or healthy individuals (HC) and treated with or without Belzutifan (50 nM) in five different time points: T0, 12, 24, 36, and 48 hours. Data are shown as means ± SD for HC (n=8), HC+Belzutifan (n=6), PCS (n=10), PCS+Belzutifan (n=9). A p-value of <0.05 was considered statistically significant. Adjusted p values were determined by Tukey multiple comparisons tests in 2-way ANOVA

## Discussion

As evidence arises that persistent viral SARS-CoV-2 RNA and proteins may sustain ongoing symptoms after acute infection the aim of this work was to study the effect of S1 antigen on endothelial function [43]. We were able to demonstrate that S1-driven activation of HIF pathways leads to ED, characterized by increased oxidative stress and barrier dysfunction. Treatment with Belzutifan effectively restored endothelial integrity.

In our PCS cohort VEGF levels were significantly elevated, consistent with reports of elevated ED markers in PCS [44–46]. Studies report higher Anti-S1 IgG levels and/or viral coinfections in PCS patients maintaining chronic inflammation, autoimmunity, and persistent endothelial activation [47–49]. In our cohort Anti-S1 was positively correlated with VEGF, potentially linking persistent S1 and ED. Additionally, in our *in vitro* experiments S1 treatment of HREC was capable to induce VEGF mRNA expression in HREC. Endotheliitis in acute SARS-CoV-2 infection is contributing to vascular complications. We were surprised that S1 alone was not capable to induce the expression of inflammatory markers or NF-kB in HREC. Activation of inflammatory pathways in EC seems to be depended on higher S1 concentrations and/or indirectly via the activation of immune cells. As we wanted to mimic a chronic, underlying infection, we used a low concentration of S1 compared to other publications (100 ng/mL vs. 750000 ng/mL) [50–52]. Therefore, we propose that VEGF expression in HREC is driven by S1-induced HIF-1α and HIF-2α activation, rather than the NF-kB. S1 induced significant HIF-1α nuclear translocation peaking at 8 hours post-treatment with a sharply decrease after 24 hours, while HIF-2α remained active beyond 72 hours, indicating a shift from HIF-1α to HIF-2α prolonged hypoxia [27, 28, 53]. Interestingly, supporting our hypothesis, a recent analysis of the plasma proteome of PCS patients revealed enrichment of HIF signaling pathways [54]. The observed down-regulation of IL-8 mRNA by S1 may be due to the activation of HIF-1α, which is known to inhibit NF-E2-related factor (Nrf2), a transcription factor that modulates IL-8 in EC [55, 56] S1 has been hypothesized to interact with Hb, disrupting its function, reducing oxygen transport, and inducing hypoxia [57, 58]. We also found a weak albeit significant association between higher Anti-S1 IgG levels and low Hb in PCS patients. Hb and Hct were negatively correlated with PCS severity, which suggests a potential role of chronic hypoxia in PCS symptom development. We observed significantly higher EPO levels in PCS patients, which further underscores the presence of chronic hypoxia in these individuals. This aligns with a study reporting persistent iron dysregulation and inflammatory anemia associated with PCS and studies reporting higher EPO levels in PCS patients [45, 59].

In our *in vitro* experiments we demonstrated that S1 induces cellular and mitochondrial ROS in HREC. Recent studies have shown that S1 alone can promote blood brain barrier disruption and that persistent S1 might contribute to cognitive impairment in PCS [60, 61]. Following S1 treatment, EC exhibited an elongated morphology, characterized by the disappearance of the cortical actin rim and the formation of prominent actin stress fibers across the cell body. VE-Cadherin and Claudin-5 were diminished in the cell–cell contacts of S1-treated HREC. TEER significantly decreased in S1-treated cells, confirming S1-induced monolayer permeability and barrier compromise.

Similarly, we showed that EC exposed to plasma from PCS patients exhibited a notable increase in cellular and mitochondrial ROS production and a pronounced reduction of NO synthesis. TEER from monolayers exposed to PCS-plasma showed a visible however non-significant endothelial barrier disruption compared to HC-plasma.

We propose that microvascular impairement in PCS might be a consequence of persisting viral componentes, such as S1, resulting in ongoing chronic inflammation, inflammatory anemia, and potential complement-mediated tissue injury [47, 48, 62].

These mechanisms are not SARS-CoV-2 specific and have been observed in other chronic viral infections like CMV, EBV, and HHV6 [63, 64]. The overlap between PCS and ME/CFS is well described, with similar findings of decreased NO production in EC exposed to ME/CFS patient plasma and associations between activation of HIF-2α and ME/CSF [59, 65–67].

To date, there is a lack of available therapies and pharmacological trials, leaving a gap in effective treatment for patients suffering from PCS [68]. Here, we were able to show, that Belzutifan significantly restored barrier function in HREC treated with S1 or PCS plasma. We hypothesize that HIF-2α inhibition reduces stress fiber formation and promotes junction protein expression leading to a normal barrier function [71, 72].

Strengths of our study include the well-characterised PCS cohort with patient reported outcome measures in combination with in vitro cell culture experiments with patients samples.

Nevertheless, this study has some limitations. The observed effect in our *in vitro* experiments might be overinterpreted as we specifically used the plasma of PCS patients with the highest severity score. We lack data from a cohort of individuals who have recovered from COVID-19 infection or have been vaccinated. This study provides exploratory-driven results. Although, if possible, we tried to correct for potential confounders, findings must be validated in larger, independent cohorts.

Taken together this study extends the knowledge of persistent ED as a hallmark of PCS. We suggest a hypoxia-induced, HIF-signaling-mediated pathway as a driver of sustained endotheliopathy in PCS patients. Upstream events include elevated ROS production and decreased NO levels, which activate HIF-1α and HIF-2α, leading to endothelial barrier disruption. These results potentially explain current findings of blood-brain barrier disruption in PCS-associated cognitive impairment and add to the pathomechanistic understanding of PCS. While the exact trigger of chronic hypoxia in PCS patients remains unclear, we propose that persistent changes in erythropoiesis, driven by residual S1 and/or reactivation of other viral entities might cause chronic hypoxia in PCS patients. Our findings indicate that employing Belzutifan, a HIF-2α pathway inhibitor, may restore endothelial function. Ultimately large-scaled studies need to further clarify the exact trigger of HIF-mediated ED in PCS. However, if confirmed, Belzutifan could be a very promising therapeutic approach in treating PCS.

## Data Availability

All data produced in the present study are available upon reasonable request to the authors

## Nonstandard Abbreviations and Acronyms

ACE2: angiotensin-converting enzyme-2
C19-YRS: COVID-19 Yorkshire Rehabilitation Scale
CoCl2: Cobalt chloride
CXCL1: C-X-C motif chemokine ligand 1
CXCL10: C-X-C motif chemokine ligand 10
EC: Endothelial cells
ECIS: Electrical cell-substrate impedance sensing
ED: Endothelial dysfunction
EPO: Erythropoietin
FSS: Fatigue Severity Scale
Hb: Hemoglobin
HC: Healthy control
Hct: Hematocrit
HIF: Hypoxia inducible factor
HREC: Human retinal endothelial cells
ICAM-1: Intercellular adhesion molecule 1
IL-6: Interleukin-6
IL-8: Interleukin 8
LPS: Lipopolysaccharide
MCP-1: Monocyte Chemoattractant Protein-1
ME/CFS: Myalgic encephalomyelitis/chronic fatigue syndrome
Nrf2: NF-E2-related factor
NO: Nitric oxide
PCS: Post-COVID Syndrome
PHQ-9: Patient Health Questionnaire 9
PROM: Patient reported-outcome measures
ROS: Reactive oxygen species
RT-PCR: Reverse transcription-polymerase chain reaction
S1: Spike protein 1
SARS-CoV-2: Severe-Acute-Respiratory-Syndrome Coronavirus 2
SD: Standard deviation
TEER: Transendothelial electric resistance
TNF: Tumor necrosis factor
VEGF: Vascular endothelial growth factor

## Acknowledgements

The authors thank for the technical assistance of Sandra Haderer and Alina Schmidt.

